# Assessment of ten-year cardiovascular disease risk in Bangladesh: A comparative analysis of laboratory and non-laboratory-based WHO risk prediction models

**DOI:** 10.1101/2024.07.04.24309982

**Authors:** Md Mostafa Monower, Shehab Uddin Al Abid, Mahfuzur Rahman Bhuiyan, Mohammad Abdullah Al Mamun, Sohel Reza Choudhury

## Abstract

**Objectives:** This study aimed to estimate the 10-year cardiovascular diseases (CVD) risk in Bangladeshi adults using updated 2019 WHO laboratory-based and non-laboratory-based models, identify key CVD risk determinants, and assess the agreement between two models.

**Methods:** We conducted a cross-sectional analysis of nationally representative Bangladesh STEPS 2018 survey data, involving 2767 adults aged 40-69 years without prior CVDs, using both versions of the updated 2019 WHO 10-year CVD risk prediction models. Logistic regression identified determinants and model agreement was assessed using Bland-Altman Plot, concordance, and Kappa statistics with STATA version 17.

**Results:** In the laboratory-based model, 10.32% (95% CI: 8.78-12.11) of the population was at elevated risk (≥10%), while the non-laboratory-based model showed 9.25% (95% CI: 7.7-11.08). Males had significantly higher risk than females in both models (p<0.0001). Elevated 10-year CVD risk was associated with age increments of five years (OR=4.29, 95% CI: 3.48-5.30), being female (OR=0.31, 95% CI: 0.18-0.53), urban residence (OR=1.82, 95% CI: 1.20-2.74), residing in Dhaka division (OR=2.44, 95% CI: 1.26-4.73), and higher education (OR=2.42, 95% CI: 1.06-5.51). Key metabolic factors included high waist-hip ratio (OR=2.08, 95% CI: 1.22-3.52) and elevated triglycerides (OR=2.70, 95% CI: 1.65-4.41). Comparing models, Bland-Altman plots showed limit of agreement (-3.16, 3.52) with mean difference of 0.18, proportional bias (males: y=-0.43 + 1.11x; females: y=-0.43 + 1.16x), Lin’s Concordance Correlation Coefficient (LCCC) 0.90 (95% CI: 0.89-0.91), and a Kappa statistic of 0.69 (95%CI: 0.67-0.72).

**Conclusion:** A substantial portion of the adult Bangladeshi population is at elevated risk of a cardiovascular event within the next decade, with males at higher risk per both models. Sociodemographic, and metabolic factors indicating potential target groups for intervention. The non-laboratory-based model generally shows agreement with the laboratory-based model and can serve as an alternative in settings with limited laboratory resources, though caution is advised for high-risk individuals.

## Introduction

Cardiovascular diseases (CVDs) stand as the leading cause of global mortality, accounting for an estimated 17.9 million lives annually and constituting about 32% of all deaths worldwide.[1] The majority of these fatalities occur in low and middle-income countries (LMICs),[1] where challenges such as inadequate healthcare infrastructure, limited access to essential services, and low health literacy intensify the impact of these burden.[2], [3] Bangladesh, characterized as an LMIC, is experiencing a rapidly growing burden of CVDs, propelled by demographic shifts and lifestyle changes.[4]

Early identification of at-risk populations is one of the effective strategies to prevent CVDs.[5], [6] In this context, the World Health Organization’s (WHO) updated risk prediction charts can serve as a valuable tool,[7] which can identify individuals who would benefit most from preventive interventions such as statin therapy.[8] These charts, revised in 2019, estimate the 10-year risk of a major cardiovascular event using factors such as age, gender, blood pressure, smoking status, cholesterol level, and presence of diabetes.[9] The adaptations of these models for 21 different global regions improve CVD risk prediction accuracy by accommodating variations in risk factor levels and disease incidences across diverse settings.[9] Additionally, these new models incorporate both fatal and non-fatal cardiovascular outcomes, providing a more extensive assessment of CVD risk compared to models that only predict mortality.[9]

Bangladesh aims to achieve a 25% reduction in premature mortality from non-communicable diseases (NCDs) by 2025, as part of its commitment to reducing CVD impact,[10] continuing and expanding upon the targets set in earlier plans.[11] For resource limited settings like Bangladesh, the revised WHO 2019 models offer a viable strategy for assessing CVD risk and facilitating targeted interventions such as lifestyle modifications and pharmacotherapy.[6], [12] These models are available in two forms: a laboratory-based version that includes biochemical markers like total cholesterol and blood glucose, and a non-laboratory-based version that relies on clinical and demographic information alone.[8], [9]

Evaluation of these models within the Bangladeshi context, the 2018 Bangladesh STEPS survey, provides a unique opportunity through its collected comprehensive data on non-communicable disease risk factors.[13] By identifying potential target groups for intervention, and by evaluating whether simpler, non-laboratory methods can substitute for lab-based assessments, this study could markedly influence public health policies and patient management strategies, not only in Bangladesh but also as an example for similar global settings. In this context, this study aimed to estimate the prevalence of the 10-year cardiovascular disease risk using both laboratory and non-laboratory-based models, identify the key determinants of CVD risk, particularly using the laboratory-based model, and assess the agreement between the two models.

## Methodology

### Study design setting and sampling

This study used data from the 2018 Bangladesh STEPS survey, focusing on non-communicable diseases (NCDs). Conducted from February to May 2018, the survey employed a cross-sectional design with two-stage stratified cluster sampling. Of 496 initial primary sampling units (PSUs), 495 were selected from urban and rural sectors across all eight divisions. In each PSU, 20 households were systematically chosen, and one adult was randomly selected per household. A total of 8,185 individuals completed the interview, 7,208 completed physical measurements, and 7,056 provided blood samples.[13], [14] After applying inclusion criteria for age (40-69 years) and exclusion criteria for previous CVDs and missing data, the study included 2,767 participants from 493 PSUs with complete datasets across all measured variables (**S1 Fig**). Data completeness was paramount; therefore, observations with missing values were excluded from the analysis. We utilized a complete-case analysis approach, which was justified due to the minimal impact of missing data: only 0.8% (n=22) of the data were missing (SBP=1, WHR=4, BMI=2, and education level=15). This approach was not expected to introduce any substantial bias, as the percentages of missing data for these variables were very low.

### Data collection

Data collection adhered to the WHO STEPS methodology (Version 3.2) and was conducted in three phases. STEP-1 involved structured interviews to gather demographic and lifestyle risk factors such as residence, education, occupation, diet, physical activity, and tobacco use, acknowledging that some degree of recall bias may still be present in self-reported data. STEP-2 included physical measurements and blood pressure assessments. In the final phase, STEP-3, blood samples were collected under aseptic conditions to ensure data accuracy and reliability. Participants were required to fast for 12 hours prior to sample collection, while individuals with diabetes were advised to bring their medication and take it immediately after the sample was collected. The detailed methodological procedure was described in the 2018 STEPS Bangladesh report.[13], [14]

### Blood pressure measurements

Blood pressure was measured using “BP–BOSO–Medicus Control” digital monitor, which was validated by the German Hypertension League.[15] Participants were asked to rest for 15 minutes with uncrossed legs before measurements. Three consecutive readings were taken at three-minute intervals, and the mean of the last two was computed for analysis.[13]

### Blood sample collection

Participants fasted overnight before their appointment for blood sample collection. During the visit, 5 ml of blood was drawn: 2 ml was allocated for serum glucose testing in a Fluoride-oxalate vacutainer, and 3 ml was used for a lipid profile analysis in a standard tube. Samples were processed and transported to the NIPSOM Lab within 24 hours, with stringent cold chain.[13]

### Biochemical analysis

Blood glucose levels were assessed using a Human® kit from Germany. TG levels were measured using Elitech® kits with controls from Humatrol/serodos®. Cholesterol levels were determined using an Elitech® kit with Humatrol® controls from Germany. All assays were conducted at the NIPSOM laboratory using a biochemistry auto analyzer (Selectra Pro M).[13] ***Estimation of 10-year CVD risk***

To estimate the 10-year risk of CVDs, we utilized the updated 2019 WHO 10-year CVD risk prediction models, which were tailored for 21 WHO epidemiological subregions and offered two versions per region-one requiring laboratory results and another not.[8], [9] Given the wide-ranging data collection in this dataset, including cholesterol measurements, we employed both the risk models, utilizing a STATA command developed by the authors of the revised WHO 2019 10-year CVD risk prediction models and tailored for Bangladesh.[16] The risk estimation was designed to calculate on various factors.

- Age: Grouped as 40–44, 45–49, 50–54, 55–59, 60–64, and 65–69 years.
- Sex: Analyzed by male and female categories.
- Smoking status: Classified participants as smokers or non-smokers.
- Systolic blood pressure (SBP): Categories included <120 mm Hg, 120–139 mm Hg, 140–159 mm Hg, 160–179 mm Hg, and ≥180 mm Hg.
- Body mass index (BMI): BMI was calculated by dividing a respondent’s weight in kilograms (kg) by the square of their height in meters (m²); <20, 20–24, 25–29, 30– 34, and ≥35 kg/m².
- Total cholesterol: Laboratory measured blood cholesterol level (mmol/L).
- Diabetes: Defined by a fasting blood glucose level >126 mg/dl or current use of antidiabetic medication; categorized as present or absent.

The CVD risk levels were categorized into five groups indicating the percentage risk of likelihood of experiencing a fatal or non-fatal major cardiovascular event, such as myocardial infarction or stroke, over the next 10 years: <5%, 5% to <10%, 10% to <20%, 20% to <30%, and ≥30%.[8]

### Explanatory variables

In this study, the outcome variable was defined based on the total CVD risk score, with a score of ≥10% classified as elevated CVD risk and a score of <10% classified as lower CVD risk.[17] Potential factors associated with elevated CVD risk among the Bangladeshi population were identified through a literature review and the availability of data from this survey.[4], [18], [19] Sociodemographic variables considered included residence type (rural or urban) and geographic division of the country (Barisal, Chittagong, Dhaka, Khulna, Mymensingh, Rajshahi, Rangpur, and Sylhet), as well as educational attainment (no formal education, primary, secondary, and more than secondary). Metabolic factors included waist hip ratio and blood triglyceride level. Central obesity was determined using waist-hip-ratio thresholds of ≥0.90 for males and ≥0.85 for females.[20] A triglyceride level of ≥150 mg/dl was classified as elevated.

### Statistical analysis

Descriptive statistics were used to summarize the WHO 10-year CVD risk, stratified by gender, presenting weighted percent prevalence and 95% confidence intervals. Pearson’s chi-squared test was conducted to assess the significance of differences. Variables estimating CVD risk and associated covariates were detailed by frequency, percentage, or medians with interquartile ranges (25th to 75th percentiles) as appropriate, as shown in **S1 & S2 Table**. Sampling weights were incorporated to estimate prevalence more accurately, ensuring a representative analysis of the study population.

To assess the influence of potential confounders on the association between 10-year lab-based CVD risk and its determinants, a stepwise design adjusted logistic regression was performed. This model adjusted for age, sex, and socio-demographic variables (urban/rural residence, division, education level), and metabolic risk factors (waist-hip ratio and triglyceride level). Variables traditionally used for estimating CVD risk such as smoking status, systolic blood pressure, history of diabetes, total cholesterol, and body mass index were excluded from the model, with the exception of non-modifiable factors like age and sex. Outcomes were dichotomized, and independent variables were included methodically based on literature recommendations. The associations were illustrated using odds ratios (ORs) with 95% confidence intervals (CIs). All statistical analyses were two-tailed, with a significance level set at p<0.05 for main effects and p<0.001 for interaction effects. Multicollinearity was assessed using the variance inflation factor (VIF), with a threshold of five indicating significant multicollinearity.

The agreement between lab-based and non-lab-based risk charts was evaluated using Bland-Altman plots and Lin’s concordance coefficient correlation (LCCC) for continuous CVD risk scores, and Cohen’s kappa for categorical CVD risk classifications. The Bland-Altman method plotted the difference between lab-based and non-lab-based risk scores against their average, and limits of agreement were calculated as the mean difference ± 1.96 standard deviations (SD) of the mean difference. The agreement between the laboratory-based and non-laboratory-based cardiovascular risk predictions was further assessed using Lin’s Concordance Correlation Coefficient (LCCC), which ranges from -1 to 1, with 1 indicating perfect concordance. Cohen’s kappa was employed to measure the categorical agreement between the two risk charts, categorizing kappa values as follows: less than 0 indicating no agreement; 0.01 to 0.20 as slight agreement; 0.21 to 0.40 as fair agreement; 0.41 to 0.60 as moderate agreement; 0.61 to 0.80 as substantial agreement; and 0.81 to 1.00 as almost perfect agreement. Data preparation and analysis were conducted using STATA version 17, ensuring rigorous statistical examination and reproducibility of the results.

### Ethical consideration

The protocol for the 2018 Bangladesh STEPS study was approved by the Bangladesh Medical Research Council (BMRC). Before conducting interviews and collecting specimens, informed written consent was obtained from all participants. Furthermore, in March 2023, the NCD Microdata Repository of the World Health Organization (WHO) granted permission to access and use the dataset. Since this research involves secondary data that was collected with prior ethical approval and participant consent, no additional ethical clearance or further consent for publication was required. Our study complies with the ethical guidelines for the analysis of secondary data.

## Results

The results presented in **Table 1** indicate estimated prevalence of 10-year cardiovascular disease (CVD) risk, stratified by gender and assessed using both laboratory-based and non-laboratory-based revised WHO models. In the laboratory-based model, more than two-third portion of the population was categorized at very low risk (<5%), but higher risk categories also indicated important observations. Specifically, the prevalence of low risk (5%-<10%), moderate risk (10%-<20%), high risk (20%-<30%), and very high risk (≥30%) were 26.62% (95% CI: 24.35-29.03), 9.49% (95% CI: 7.96-11.27), 0.81% (95% CI: 0.42-1.58), and 0.03% (95% CI: 0.01-0.12), respectively. Notably, males exhibited significantly higher prevalence across these elevated risk categories compared to females (p<0.0001).

**Table 1:**
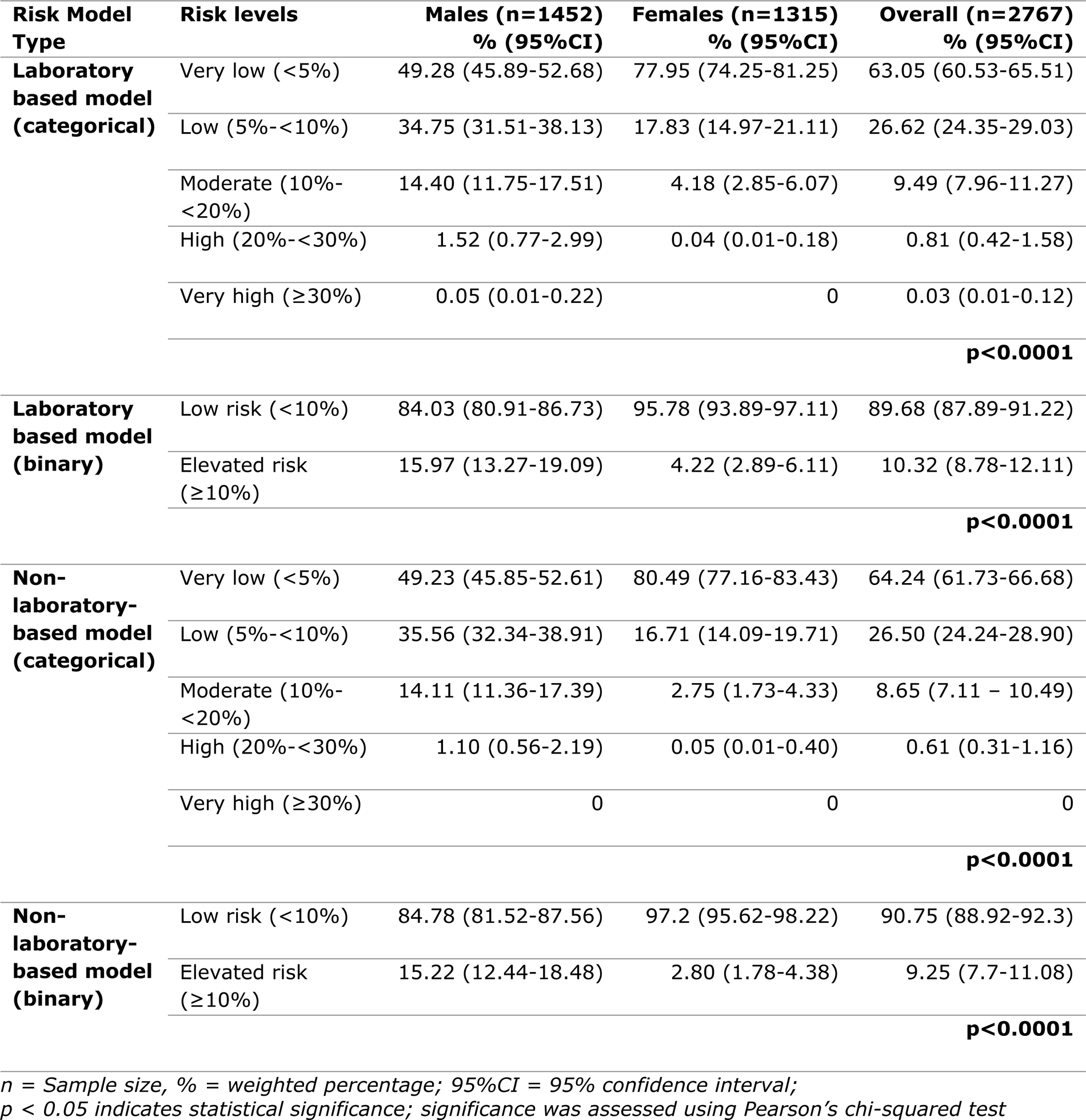
Estimated prevalence of WHO 10-year CVD risk by gender.

Similarly, the non-laboratory-based model showed comparable patterns. The prevalence for low risk was 26.50% (95% CI: 24.24-28.90), for moderate risk was 8.65% (95% CI: 7.11-10.49), and for high risk was 0.61% (95% CI: 0.31-1.16). No individuals were recorded at the very high-risk level. This model also indicated clear gender disparities, with males consistently demonstrating higher risk levels than females (p<0.0001).

When categorizing risk into broader categories (**Table 1**), in the laboratory-based model, 10.32% (95% CI: 8.78-12.11) of the overall population was identified at elevated risk (≥10%), with a significant disparity between genders. Specifically, 15.97% (95% CI: 13.27-19.09) of males were at elevated risk, contrasted with only 4.22% (95% CI: 2.89-6.11) of females showing the same level of risk. Similarly, the non-laboratory-based model demonstrated that 9.25% (95% CI: 7.7-11.08) of participants were at elevated risk, with males again showing a higher risk prevalence of 15.22% (95% CI: 12.44-18.48) compared to 2.80% (95% CI: 1.78-4.38) among females. The marked gender differences in elevated risk were statistically significant in both models (p<0.0001), emphasizing the robustness of these findings and indicating a significant gender-based divergence in CVD risk across the Bangladeshi population.

Age-sex adjusted and multivariate adjusted forest plot (**Fig 1**) identified several associations between sociodemographic and metabolic factors with the elevated (≥10%) 10-year CVD risk. In the multivariate adjusted model, age showed a strong association, with each five-year increase in age nearly quadruples the risk (OR = 4.29, 95% CI: 3.48-5.30). There was a notable difference by gender; females had lower odds compared to males (OR = 0.31, 95% CI: 0.18-0.53). Urban residency was associated with greater odds compared to rural residency (OR = 1.82, 95% CI: 1.20-2.74). Among divisions, Dhaka showed a higher association (OR = 2.44, 95% CI: 1.26-4.73) relative to Barisal. Higher educational attainment correlated with increased odds, with those having more than a secondary education showing the highest odds (OR = 2.42, 95% CI: 1.06-5.51). Metabolic factors such as a substantially high waist-hip ratio (OR = 2.08, 95% CI: 1.22-3.52) and elevated triglyceride levels (OR = 2.70, 95% CI: 1.65-4.41) were also associated with higher odds. The comparison between age-sex adjusted model and multivariate adjusted model demonstrated minor variations of odds ratios (ORs), indicating stable estimates upon the inclusion of additional sociodemographic and metabolic factors.

**Fig 1:**
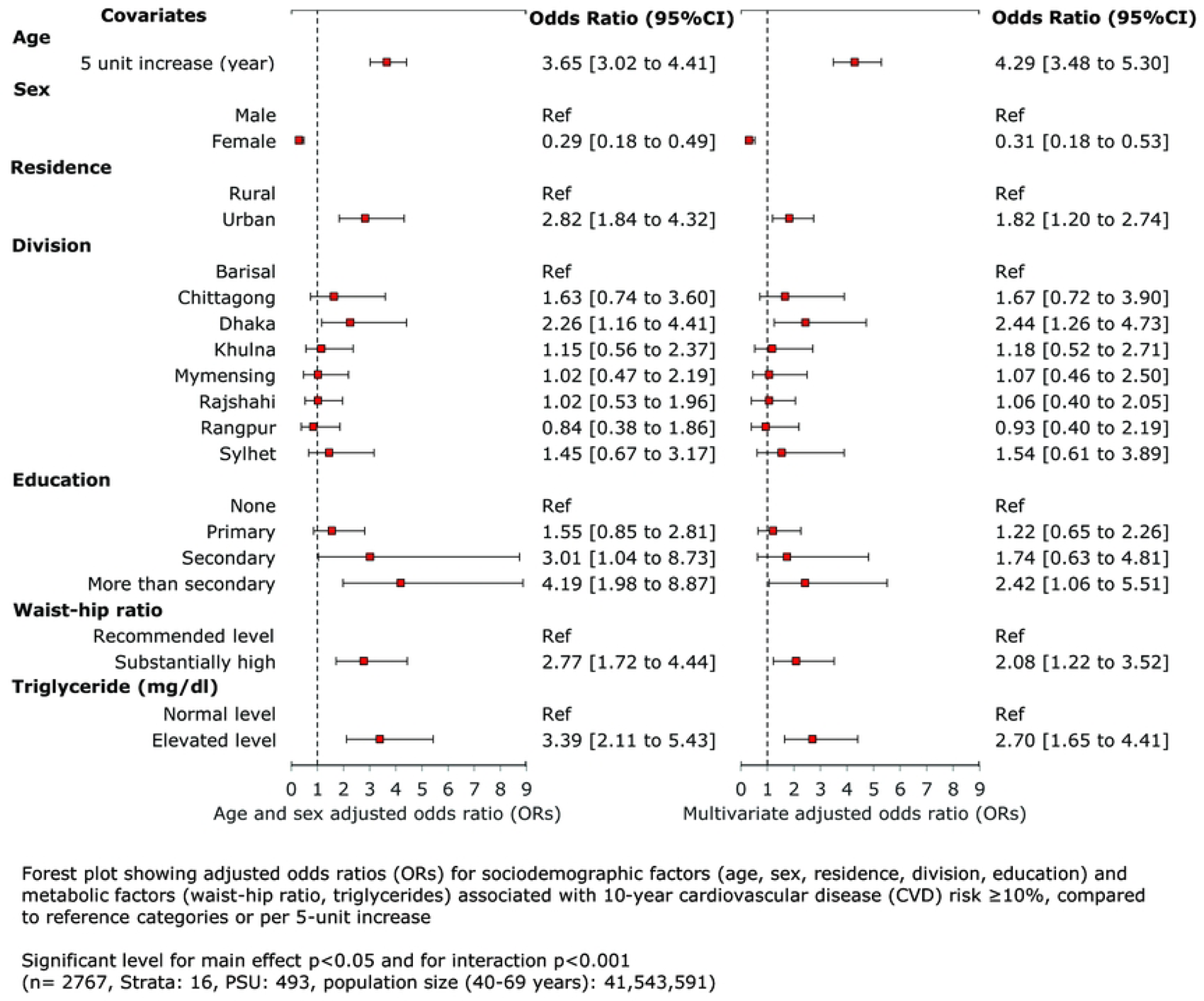
Association of laboratory based 10-year CVD risk (≥10%) and its determinants

### Limits of agreement

The Bland-Altman plots (**Fig 2**) for the overall population and both sexes demonstrated the level of agreement between laboratory-based and non-laboratory-based CVD risk scores. The average of the two methods was plotted against their difference, with the mean difference indicated by a purple line and the limits of agreement by dotted red lines at +1.96 and -1.96 standard deviations from the mean difference. For the overall population, the mean difference of 0.18 indicated that, on average, laboratory-based scores were slightly higher than non-laboratory-based scores. The 95% limits of agreement, ranging from -3.16 to 3.52, covered most differences and suggested reasonable agreement, considering clinical importance. For males, the mean difference was 0.22 with limits of agreement from -3.51 to 3.96, showing substantial variability despite general agreement. For females, the mean difference was lower at 0.13, with narrower limits of agreement from -2.71 to 2.97, indicating slightly better consistency than in males.

**Fig 2:**
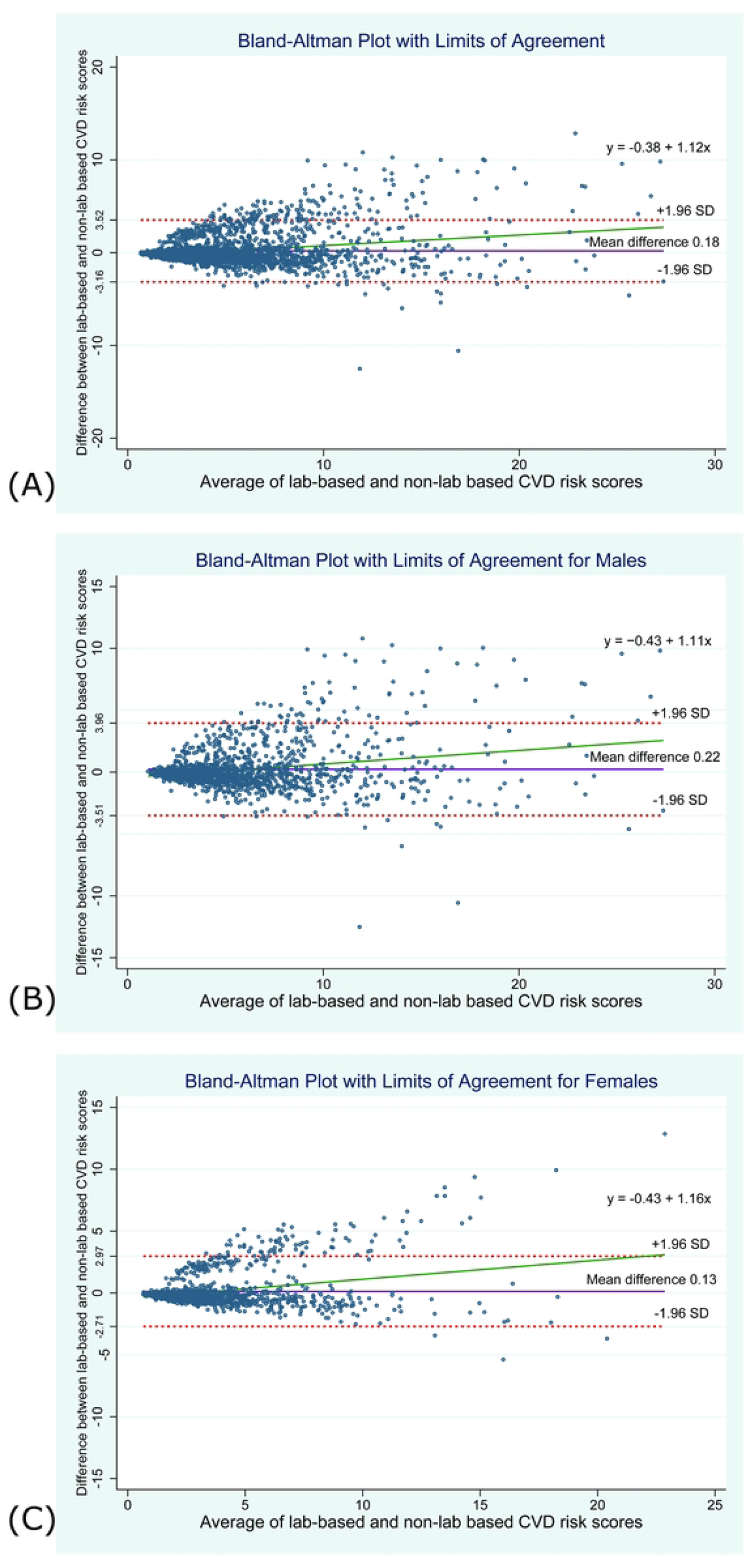
Bland-Altman plots level of agreement between laboratory-based and non-laboratory-based WHO 10-year CVD risk scores (A) Overall population (B) Among males (C) Among females

Across all plots, there was a noticeable trend indicating that the discrepancy between the laboratory-based and non-laboratory-based methods increased as the average CVD risk scores rose. This proportional bias, highlighted by the regression equations (males: y=-0.43 + 1.11x; females: y= -0.43 + 1.16x), suggested that differences widened more than proportionally with rising risk scores, which was higher in females, where the regression line’s steeper slope pointed to greater discrepancies at higher scores.

### Agreement by Lin’s Concordance Correlation Coefficient (LCCC)

The Lin’s Concordance Correlation Coefficient (LCCC) value was found to be 0.90 (95% CI: 0.89-0.91), as shown in **Table 2**, indicating a high level of agreement between the laboratory-based and non-laboratory-based 10-year CVD risk scores. Upon stratification by sex, the LCCC was calculated at 0.89 (95% CI: 0.88-0.90) for males and 0.87 (95% CI: 0.86-0.88) for females, demonstrating similarly high agreement for both genders in the 10-year CVD risk assessments.

**Table 2:** Agreement between laboratory-based and non-laboratory-based 10-Year CVD risk scores by Lin’s concordance correlation coefficient and kappa statistic.

| Measurement | Male | Female | Overall |
| --- | --- | --- | --- |
| **Lin's Concordance Correlation Coefficient (LCCC)** |  |  |  |
| **LCCC** | 0.89 (95%CI: 0.88-0.90) | 0.87 (95%CI: 0.86-0.88) | 0.90 (95%CI: 0.89-0.91) |
| **Kappa Statistic (Categorical Agreement)** |  |  |  |
| **Kappa** | 0.68 (95%CI: 0.64-0.71) | 0.66 (95%CI: 0.61-0.71) | 0.69 (95%CI: 0.67-0.72) |

*95%CI = 95% confidence interval*

### Categorical agreement

In the overall population, substantial agreement was observed (**Table 2**) between the laboratory-based and non-laboratory-based methods when the risk scores were categorized using a five-group classification system based on different levels of CVD risk. The Kappa statistic for the entire sample was 0.69 (95%CI: 0.67-0.72), indicating substantial agreement. When stratified by sex, the level of agreement varied; for males, the Kappa statistic was 0.68 (95%CI: 0.64-0.71), and for females, the Kappa statistic was slightly lower at 0.66 (95%CI: 0.61-0.71). These findings indicate that both methods generally exhibited substantial agreement across the total population and both genders, suggesting that the observed patterns are unlikely to be due to random variation.

## Discussion

This study assessed the 10-year risk of fatal and non-fatal CVD in Bangladesh using both laboratory-based and non-laboratory-based revised WHO 2019 risk prediction models, highlighting sociodemographic and metabolic factors associated CVD risk. Moreover, the findings demonstrated the agreement between the models to underscore the similarity of estimation.

The laboratory-based model identified 10.32% of the population as being at elevated risk (≥10%) (**Table 1**), slightly higher than the 9.25% identified by the non-laboratory-based model. This prevalence was lower than those reported in previous studies in Bangladesh, which reported rates from 14.9% to 27.5%.[12], [17], [21], [22] Comparative studies from other LMIC regions reported lower moderate to high CVD risk prevalence: 8.8% (laboratory-based) and 12.6% (non-laboratory-based) in Bhutan, and 8.4% (laboratory-based) and 7.5% (non-laboratory-based) in Eastern Sub-Saharan Africa.[23], [24] Over three-fifths of the population were categorized as very low risk (<5%), and about one-fifth as low risk (5%-<10%), with similar patterns in both models. Although the likelihood of cardiovascular events is generally lower in people with lower risk, it is crucial to understand that they are not entirely risk-free.[25] If effective public health measures are not implemented, cardiovascular events could still occur among those with lower risk categories, who make up the majority of the population.[26]

The low readiness index in primary care settings, such as Upazila Health Complexes (UHCs), compared to District Hospitals (DHs), indicates that the primary healthcare system is ill-equipped to handle the growing burden of CVD. Facilities that offer both diagnostic and treatment services for CVDs are limited, and only a small fraction have adequately trained staff for managing these conditions.[27] The introduction of NCD corners has increased awareness and improved care for conditions like diabetes, hypertension, and chronic obstructive pulmonary disease. However, challenges persist, such as the absence of specific guidelines, a shortage of trained personnel, inadequate laboratory facilities, insufficient logistics and medications, and poor recording and reporting systems.[28]

Among the population, the impacts of both sociodemographic and metabolic determinants on CVD risk were clearly delineated in the age-sex adjusted and multivariate adjusted models. Age was confirmed as a fundamental determinant, with each five-year increment nearly quadrupling CVD risk, mirroring global data that underscores aging as a primary risk factor for cardiovascular deterioration.[29] Gender differences were also evident, with females displaying significantly lower risk odds compared to males, aligning with international studies that suggest hormonal protections in females and generally later onset of CVD.[30] These results are consistent with global trends observed in other studies, which consistently report higher CVD risk in males across various populations.[31], [32]

Urban residency was associated with higher CVD risk compared to rural, reflecting the influence of urbanization on health through lifestyle changes such as increased sedentary behavior and dietary shifts towards processed foods.[19] Living in the Dhaka division was also associated with CVD risk, consistent with previous research identifying stress and pollution as contributing factors.[33] Surprisingly, higher educational attainment correlated with increased CVD risk, contrasting with some global findings where education typically reduces risk through improved health literacy and access to care.[34] In Bangladesh, this may highlight adverse lifestyle factors associated with higher education and socio-economic status, such as less physical activity and higher caloric intake.[19] Metabolic factors, including waist-hip ratio and elevated triglyceride levels, were strongly linked with higher CVD risk, consistent with findings from the REACTION study.[35] The robustness of these associations, demonstrated by their consistency across various model adjustments, suggests that the association are minimally influenced by confounding factors.

The Bland-Altman analysis in this study explained a general agreement between laboratory-based and non-laboratory-based CVD risk assessment methods, with minor mean differences, underscoring the role of biochemical markers in risk calculations. Furthermore, narrow limit of agreement for females indicated better consistency compared to males. These observations support existing literature that validates non-laboratory methods as reliable alternatives for CVD risk assessment, making them particularly beneficial in resource-constrained environments.[23], [36] The Lin’s Concordance Correlation Coefficient (LCCC) values demonstrated substantial agreement (overall LCCC = 0.90), with slightly higher concordance in males. These findings confirm the consistency of both assessment methods in evaluating CVD risk, supporting their use across various clinical and epidemiological settings. Similar findings of high concordance between the two WHO CVD risk assessment methods were reported in comparative studies from Bangladesh and India.[21], [37] Moreover, the Kappa statistics indicated substantial agreement across the total population and both genders using the five-group classification system. Similarly, studies conducted in North India reported lower Kappa values, whereas those in Bhutan noted higher values.[23], [37] This agreement is key for public health strategies, aiding high-risk population identification and intervention. Non-laboratory methods are beneficial for their simplicity and low resource needs.

The regression line indicated an increasing discrepancy between the two methods as average CVD risk scores rose, which was more evident in females. This trend suggests the non-laboratory method may struggle with complex risk factors at higher CVD risk levels. Despite the general agreement and validation of non-laboratory methods as cost-effective alternatives, the observed increasing discrepancies with rising CVD risk scores in both genders necessitate cautious use, particularly for higher risk assessments among female individuals, and suggest potential benefits from complementary detailed laboratory evaluations.

### Clinical and policy implication

The implications of findings from this paper are influential for both clinical practice and policy development. Clinically, non-laboratory-based CVD risk scores should be used cautiously for high-risk individuals, who may require precise evaluations using laboratory-based methods. From a policy perspective, integrating WHO risk prediction models into national health guidelines and screening programs could effectively identify at-risk populations, supporting cost-effective preventive strategies. Leveraging simpler, non-laboratory models for widespread use, particularly in resource-limited settings, can effectively reach a broader population. Training community health workers to conduct assessments and initiate preventive measures, such as providing health advice and making referrals, could further democratize access to critical health services and enhancing the health resilience of the Bangladeshi population.[8]

### Strengths and limitations

This study used both laboratory-based and non-laboratory-based WHO updated risk prediction models tailored for the Bangladeshi population, enhancing CVD risk assessment precision. Utilizing a country representative dataset ensures findings are broadly applicable across Bangladesh. Major strengths include analysis of gender variability, sociodemographic and metabolic determinants, and model comparison, enriching our understanding of CVD risk. Despite these valuable insights, the study is subject to several limitations. Its cross-sectional design limits the ability to establish causality. Self-reported data on lifestyle factors may introduce recall bias. The focus on individuals aged 40-69 does not fully represent younger adults increasingly affected by lifestyle-related health issues. Additionally, findings may not be generalizable to other countries due to variations in diet, lifestyle, and health infrastructure that are specific to Bangladesh.

## Conclusion

This study has demonstrated that a substantial portion of the adult Bangladeshi population is at elevated risk of cardiovascular event within the next decade, with males at higher risk according to both laboratory-based and non-laboratory-based models. Moreover, the findings have highlighted the association between sociodemographic and metabolic factors and cardiovascular health risk, indicating potential target groups for intervention. Notably, the non-laboratory-based model have shown substantial agreement with the laboratory-based model and can serve as an alternative in settings with limited laboratory resources, although caution is advised for high-risk individuals.

To improve the non-laboratory-based model’s accuracy at higher risk scores, future research may consider incorporating accessible predictive markers like waist circumference or detailed familial health histories. Regular validation and recalibration of both models through longitudinal studies will help maintain their relevance, particularly for younger adults. Additionally, exploring how socioeconomic and biological factors interact could deepen understanding of CVD risk disparities across different genders and socioeconomic groups. Effective intervention strategies should also be developed, leveraging community health initiatives, targeted educational programs, and mobile health technology.

## Acknowledgements

We express our gratitude to the participants of the Bangladesh STEPS 2018 survey for their invaluable contributions. This research was made possible by the dedicated efforts of the National Institute of Preventive and Social Medicine (NIPSOM) and the Ministry of Health and Family Welfare of Bangladesh. We also thank the World Health Organization for their support and for providing access to the necessary datasets.

## Author Contributions

Choudhury, Abid, and Monower conceptualized and designed the study framework. Monower conducted the statistical analysis and, along with Abid, performed the initial data interpretation. Monower drafted the initial manuscript. Bhuiyan, Mamun, Abid, Monower, and Choudhury made critical revisions for intellectual content. All authors approved the final version and agree to be accountable for all aspects of the work, ensuring questions related to accuracy or integrity are appropriately investigated and resolved.

## Patient consent for publication

Not applicable as this study did not require direct patient involvement; it utilized existing data from the Bangladesh STEPS 2018 survey.

## Data availability statement

Data for this study were obtained from the publicly accessible datasets of the Bangladesh STEPS 2018 survey, which are available via the World Health Organization NCD Microdata Repository. The data can be accessed from the repository at the following URL: https://extranet.who.int/ncdsmicrodata/index.php/home [accessed on 21 Mar 2023]. Following the repository’s instructions, the data can be downloaded.

## Supporting information

S1 Fig: Participant selection flowchart for the Study (Total n=2767)

S1 Table: Socio-demographic characteristics of the participants

S2 Table: CVD risk factors distribution of the participants

